# Early measurement of blood sST2 is a good predictor of death and poor outcomes in patients admitted for COVID-19 infection

**DOI:** 10.1101/2020.12.29.20248989

**Authors:** M Sánchez-Marteles, J Rubio-Gracia, N. Peña-Fresneda, V Garcés-Horna, B Gracia-Tello, L Martínez-Lostao, S Crespo-Aznárez, JI Pérez-Calvo, I Giménez-López

**Author notes:** Corresponding Author:* Marta Sánchez-Marteles, Internal Medicine department. University Clinical Hospital “Lozano Blesa”., San Juan Bosco Avenue, 15, 50009. Zaragoza. Spain, (Ext. 2555). Both authors contributed equally to this manuscript (M Sánchez-Marteles, J Rubio-Gracia).

## Abstract

**Importance:** Although several biomarkers have shown correlation to prognosis in COVID-19 patients, their clinical value is limited because of lack of specificity, suboptimal sensibility, or poor dynamic behavior.

**Objective:** In search of better prognostic markers in COVID-19, we hypothesized that circulating soluble ST2 (sST2) could be associated to a worse outcome, prompted by our previous knowledge of sST2 involvement in heart failure-associated lung deterioration, and by mounting evidence favoring a role of IL-33/ST2 axis in the disease.

**Design, Setting and participants:** One hundred and fifty-two patients admitted for confirmed COVID-19 infection were included in a prospective non-interventional, observational study carried out in a tertiary teaching center. Blood samples were drawn at admission, 48-72 hours later and at discharge. sST2 concentrations, and routine blood laboratory were analyzed.

**Main outcomes:** Primary end-points were admission at intensive care unit (ICU) and, mortality. Other outcomes were a need for high oxygen flow therapy (HOF) or increasing treatment at 48/72 hours.

**Results:** Median age was 57.5 years (SD: 12.8), 60.4% males. Ten per cent of patients (n=15) were derived to ICU and/or died during admission. The rest stayed hospitalized 8(IQR:6) days on average. About 34% (n=47), 38% (n=53) and 48.5% (n=66) needed HOF, up-titrate therapy or both, respectively.

Median (IQR) sST2 serum concentration (ng/mL) rose to 53.1(30.9) at admission, peaked at 48-72h (79.5[64]) and returned to admission levels at discharge (44.9[36.7]), remaining significantly elevated above healthy donor values (18.6[15.1]).

A concentration of sST2 above 58.9 ng/mL identified patients progressing to ICU admission or death. These results remained significant after multivariable analysis. The area under the receiver operating characteristics curve (AUC) of sST2 for the occurrence of end-points was 0.776 (p=0.001). Admission sST2 was higher in patients who needed up-tritate therapy.

**Conclusions and relevance:** In patients admitted for COVID-19 infection, measurement of sST2 measurement early within 24h after at admission was able to identify patients at risk of severe complications or death.

## Introduction

Coronavirus disease 2019 (COVID-19) is a worldwide pandemic caused by the novel Severe Acute Respiratory Syndrome coronavirus 2 (SARS Cov-2), an infectious disease leading to high morbidity and mortality^1,2,3^.

Clinical spectrum of COVID-19 ranges from asymptomatic patients to severe pneumoniae, with multi-organ symptoms and/or failure, including cardiological, neurological or thrombotic manifestations^4^. Regardless its broad phenotypic spectrum, prognosis depends on the development of an Acute Respiratory Distress Syndrome (ARDS), which frequently leads to the administration of High Oxygen Flow therapy (HOF) or admission at the Intensive Care Unit (ICU) and mechanical ventilation, with a very high mortality in such cases^1,3^.

As a novel disease, patientackn handling was initially based on most conspicuous clinical signs and known biomarkers of tissue damage or inflammation such us lactate dehydrogenase (LDH), interleukin 6 (IL-6), ferritin, C-reactive protein (CRP) or lymphocytes^1,3^. However, these biomarkers lack specificity, since they can rise in other critical diseases^5^, showing a poor prognostic value. For instance, initial observation of heart damage associated to COVID-19 led to investigation of troponin, whose concentrations have been found to correlate poorly with COVID-19 prognosis.^6,7,8^. Accordingly, the search for more specific, earlier and clinical useful biomarkers of poor outcomes remains crucial.

Derangement of immune response was one of the first observations in COVID-19 and still remains as a hallmark of progression into a more severe phase. Studies of lung immune cells infiltrates suggest a role for IL-33/ST2 axis^9^.

Interleukin-1 receptor-like-1 (IL1RL1), also known as suppression of tumorigenicity 2 (ST2), is a member of interleukin (IL)-1 receptor family, with two main isoforms, a transmembrane cellular (ST2L) and a circulating soluble form (sST2). ST2 is the receptor for IL-33, a cytokine released by cells in response to cell damage or stress (‘alarmin’). IL-33 binding to ST2L elicits a pleiotropic action. It is well stablished that IL-33/ST2 axis exhibits a cardioprotective role, reducing fibrosis, cardiomyocyte hypertrophy, and apoptosis and improving myocardial function^10^. For that reason, blood sST2 levels are clinically used to assess prognosis in HF. It is assumed they reflect tissue sST2 acting as a decoy receptor for IL-33 and thus reducing IL-33 cardioprotective effects. ST2 has also been associated with inflammatory phenomena^11^, specially on the vascular endothelium, and pulmonary tissues ^10,9^.

The study by *Pascual-Figal et al*. demonstrated the lung origin of sST2 in acute decompensated HF, finding a correlation between their concentrations and alveolar wall thickness. They also found sST2 is higher in patients with non-cardiogenic pulmonary edema^9,12^. Moreover, there is also convincing evidence for a role of sST2 in lung diseases such as asthma, Chronic Obstructive Pulmonary Disease (COPD) and ARDS^13,14^.

We hypothesized that sST2 circulating levels in COVID-19 patients reflects pulmonary damage and the intensity of inflammatory response elicited by SARS-CoV2, and thus could be a clinical useful biomarker able to early identify patients at higher risk of severe complications or death.

## Objectives

The main goals of our study were: 1) Determine sST2 prognostic value in COVID-19 patients. 2) Analyze sST2 dynamic behavior and establish whether it associates to clinical course. 3) Compare sST2 with other biomarkers currently used in COVID-19 (ferritin, troponin, LDH, IL-6) and 4) Analyze sST2 correlation with clinical and analytical variables.

## Material and methods

### Study design and setting

Prospective cohort study carried out at the Infectious diseases and Internal Medicine departments of a tertiary teaching center (*Hospital Clínico Universitario “Lozano Blesa”, Zaragoza, Spain*), between July and November 2020. Inclusion criteria: 1) Age ≥ 18 years. 2) Informed consent granted. 3) Confirmed diagnosis of SARS-CoV2 (COVID-19) infection by nasopharyngeal polymerase chain reaction (PCR) or specific serology (IgM and/or IgG) in the context of clinical respiratory infection. Exclusion criteria: 1) Primary admission at Intensive Care Unit. 2) Refusal to participate. 3) Functional dependence (Barthel index < 50 points). 4) Moderate/Severe cognitive impairment (Pfeiffer scale ≥ 5), 5) Advanced chronic obstructive pulmonary disease (COPD) (FEV_1_ <30%) or a history of emphysema and/or pulmonary fibrosis, 6) Active cancer.

The study complied with the fundamental guidelines of the Helsinki declaration guidelines and was evaluated and approved by Aragón’s Committee on Research Ethics (CEICA, Ref. PI20/248, May 13Th, 2020).

### Variables and definitions

Patients were studied at three specific times during COVID-19 hospitalization. 1) ‘Admission’ (first 24 hours upon admission). 2) ‘Control’ (48-72 hours later) and 3) ‘Discharge’ (last 24 hours prior to discharge). At each time point, vital signs were recorded (blood pressure, heart rate, oxygen saturation, and respiratory therapy), Kirby index (PaO_2_/FiO_2_) was estimated (ePAFI) from FiO_2_ and oxygen saturation, and patient’s dyspnea was quantified using Borg scale (range 1 [minimum] to 10 [maximum]). Routine blood laboratory data (Complete blood count -CBC-, biochemistry, coagulation, and arterial blood gasses) were recorded.

Primary outcomes were death and ICU admission. Secondary outcomes were length of stay, need for HOF, increase treatment or both of them at 48/72 hours.

### Circulating sST2 measurements

Serum sST2 concentrations were determined in 150 COVID-19 patients. Blood samples were withdrawn at admission, control and discharge. Blood was collected into clotting gel test tubes, centrifuged and serum was aliquoted and stored (Aragón’s Health System Biobank) at −80°C until analysis. Eventually, 144 admission and control, and 80 discharge samples were processed and analyzed. Serum aliquots were virus-inactivated by treatment with 1% Triton-X100. On the day of the analysis, serum was thawed and diluted 1:50 (1% Bovine Serum Albumin in Phosphate Buffered Saline). Serum concentrations of soluble ST2 were determined by sandwich enzyme-linked immuno-sorbent assay (ELISA), following manufacturer’s instructions. (Supplementary Material). A set of sera from 60 healthy donors obtained through Aragón’s Health System Biobank (BSSA) was also analyzed in this manner. These sera had been originally collected from two independent sources, and were selected to match patient cohort age and gender distribution. Random samples from COVID patients and healthy donors were re-run in independent assays to test and correct for inter-assay variability.

### Statistical analysis

Continuous variables were expressed as mean ± standard deviation (SD) or median (InterQuartile Range), as appropriate depending on normality. Categorical variables were expressed as percentages. To perform the comparative analysis between normal continuous variables, ANOVA test was used. Those variables not following normality were compared using Kruskal Wallis U test. Categorical variables were compared using the chi-square test. The analysis of the different correlations between continuous variables was carried out using the Pearson or Spearman test.

A goal of 150 inclusions was set to account for a 20% of losses, which were mostly due to the assistance pressure imposed on the clinician researchers by the pandemic situation.

In univariable and multivariable logistic regression analysis, sST2 concentration was dichotomized based on a cut-off value selected from receiver operating characteristic (ROC) curves analysis of its primary endpoint predictive value. Multivariable logistic regression model was designed to identify factors independently associated with the need of ICU transfer during admission or intra-hospital death. Candidate predictors were selected from the univariable analysis when p-value <0.200, and entered using a backward selection procedure. Age, gender and previous history of diabetes as described clinical risk factors^2^, were also included in the model. Continuous candidate variables were log-transformed (lnX) if necessary.

Confidence intervals included were 95% (95% CI), establishing statistical significance with a p lower than 0.05. Statistical analysis was carried out with Statistical Package for the Social Sciences (SPSS) version 24.0 for Windows (IBM).

## Results

### Baseline characterization of patient cohort

A total of 150 patients were sequentially included (inclusion Flow-chart depicted in Supplementary Figure 1). Participant’s mean age was 57.5±12.8 years. Sixty percent were male and 50% had bilateral pneumoniae. Baseline characteristics are summarized in Table 1.

**Table 1:** Baseline characteristics according to sST-2 concentrations (tertiles) at admission.

| Variable | TOTAL | P<25 | P25 to 75 | P >75 | P-Value |
| --- | --- | --- | --- | --- | --- |
| Total size (N) | 144 | 36 | 73 | 35 |  |
| Age (years) | 57.5 ± 12.8 | 58.5 ± 12.3 | 57.2 ± 13.6 | 57.1 ± 12.0 | 0.878 |
| Gender-Male (n[%]) | 87 (60.4) | 17 (47.2) | 44 (60.3) | 26 (74.3) | 0.066 |
| Duration of symptom (days) | 6.5 ± 3.3 | 7.0 ± 3.6 | 6.3 ± 3.3 | 6.3 ± 2.7 | 0.486 |
| Time until COVID confirmation (Days) | 3 (7) | 3 (7) | 3 (6) | 3 (6) | 0.492 |
| Comorbidities (n[%]): |  |  |  |  |  |
| • Hypertension | 54 (37.5) | 15 (41.7) | 26 (35.6) | 13 (37.1) | 0.827 |
| • Heart failure | 4 (2.8) | 1 (2.8) | 2 (2.8) | 1 (2.9) | 1.000 |
| • Dyslipidemia | 42 (29.2) | 9 (25.0) | 19 (26.0) | 14 (40.0) | 0.267 |
| • Coronary artery disease | 5 (3.5) | 1 (2.8) | 3 (4.1) | 1 (2.9) | 0.914 |
| • Diabetes | 25 (17.4) | 6 (16.7) | 9 (12.3) | 10 (28.6) | 0.113 |
| • History of smoking | 48 (33.6) | 10 (27.8) | 22 (30.6) | 16 (45.7) | 0.207 |
| • COPD/Asthma | 16 (11.1) | 3 (8.3) | 10 (13.7) | 3 (8.6) | 0.605 |
| • Atrial/flutter fibrillation | 5 (3.6) | 1 (2.9) | 3 (4.3) | 1 (2.9) | 0.901 |
| • CKD | 7 (4.9) | 3 (8.3) | 2 (2.7) | 2 (5.7) | 0.427 |
| Clinical variables |  |  |  |  |  |
| • BMI (Kgs/m2) | 28.9 (6.4) | 27.5 (.5) | 28.7 (5.7) | 30.0 (6.0) | 0.434 |
| • SBP (mmHg) | 126.9 ± 16.7 | 132.5 ± 14.4 | 125.2 ± 15.7 | 124.6 ± 17.0 | 0.066 |
| • DBP (mmHg) | 77.2 ± 10.9 | 80.7 ± 11.6 | 77.0 ± 10.3 | 74.2 ± 10.6 | 0.051 |
| • HR (bpm) | 80.9 ± 12.8 | 80.9 ± 12.1 | 81.0 ± 12.8 | 80.5 ± 13.7 | 0.980 |
| • Estimated PAFI (mmHg) | 367 (92) | 429 (101) | 403 (99) | 341 (108) | < 0.001 |
| • Borg scale for dyspnea (points) | 4 (6) | 3 (6) | 5 (5) | 4 (6) | 0.486 |
| Laboratory: |  |  |  |  |  |
| • Urea (mg/dL) | 33 (19) | 38 (16) | 32 (18) | 32 (20) | 0.069 |
| • Creatinine (mg/dL) | 0.94 (0.29) | 0.91 (0.27) | 0.88 (0.29) | 0.92 (0.36) | 0.318 |

| Variable (Continue) | TOTAL | P<25 | P25 to 75 | P >75 | P-Value |
| --- | --- | --- | --- | --- | --- |
| Laboratory: |  |  |  |  |  |
| • Aspartate aminotransferase (U/L) | 37 (27) | 30 (16) | 38 (31) | 41 (24) | < 0.001 |
| • Alanine aminotransferase (U/L) | 31 (28) | 23 (23) | 32 (43) | 33 (18) | 0.006 |
| • Creatin phosphokinase (U/L) | 94 (92) | 71 (80) | 98 (92) | 116 (143) | 0.007 |
| • Lactate dehydrogenase (U/L) | 306 (145) | 267 (70) | 310 (106) | 398 (208) | < 0.001 |
| • C-Reactive Protein (mg/L) | 63 (81) | 46 (63) | 59 (68) | 112 (137) | 0.005 |
| • Ferritin (ng/mL) | 707 (908) | 619 (838) | 676 (813) | 1338 (1061) | 0.007 |
| • Hemoglobin (g/dL) | 14.2 ± 1.5 | 14.2 ± 1.3 | 14.3 ± 1.6 | 14.1 ± 1.4 | 0.234 |
| • Total leucocytes (x 1000) | 5.6 (3.1) | 5.2 (2.1) | 5.4 (3.4) | 6.1 (3.4) | 0.251 |
| • Total lymphocytes (x 1000) | 0.9 (0.7) | 1.0 (0.5) | 0.9 (0.6) | 0.6 (0.6) | 0.021 |
| • D-Dimer (ng/mL) | 688 (633) | 719 (856) | 625 (502) | 976 (830) | 0.030 |
| • Fibrinogen (mg/dL) | 775 (208) | 739 (257) | 761 (200) | 811 (252) | 0.011 |
| • Interleukine-6 (pg/mL) | 40 (30) | 26.8 (32.4) | 42.3 (26.2) | 50.0 (24.3) | 0.008 |
| Chest X-Ray (n[%]): |  |  |  |  | 0.222 |
| • Normal | 25 (17.9) | 8 (23.5) | 13 (18.3) | 4 (11.4) |  |
| • Unilateral pneumoniae | 35 (25.0) | 9 (26.5) | 13 (18.3) | 13 (37.1) |  |
| • Bilateral pneumoniae | 80 (57.1) | 17 (50.0) | 45 (63.4) | 18 (51.4) |  |
| Baseline therapies (n[%]) |  |  |  |  |  |
| • Colchicine | 10 (6.9) | 2 (5.6) | 5 (6.8) | 3 (8.6) | 0.882 |
| • Plasma | 1 (0.7) | 0 (0.0) | 1 (1.4) | 0 (0.0) | 0.613 |
| • Remdesivir | 46 (31.9) | 9 (25.0) | 23 (31.5) | 14 (40.0) | 0.397 |
| • Systemic corticosteroids | 113 (78.5) | 26 (72.2) | 56 (76.8) | 31 (88.6) | 0.214 |
| • Medium dose of corticosteroids (Dexametasone [mg]) | 6 (3) | 6 (3) | 6 (3) | 6 (3) | 1.000 |
| • Low molecular weight heparin | 138 (95.8) | 34 (94.4) | 70 (95.9) | 34 (97.2) | 0.488 |

Variables are expressed as mean ± standard deviation or median (Interquartile range) BMI: Body Mass Index; CKD: Chronic Kidney Disease (estimated glomerular filtration rate < 60 mL/min/173m2 CKD-EPI-Creatinine method); COPD: Chronic Obstructive Pulmonary Disease; DBP: Diastolic Blood Pressure; HR: Heart Rate; bpm: beats per minute; SBP: Systolic Blood Pressure.

### sST2 concentrations

Serum concentrations of sST2 were determined in 150 COVID-19 patients and 60 age-and gender-matched healthy donors.

Median serum sST2 values in healthy donors were 18.6(15) ng/mL. We found no differences across age groups (Supplementary Figure 2), but sST2 concentrations were significantly higher in female donors (female 22.3[13] vs. male 15.7[11] ng/mL, p=0.024); Supplementary Figure 3.

A total of 369 COVID-19 blood samples were collected and analyzed for sST2 concentrations (144 at admission, 145 at control time and 80 at discharge). Admission serum sST2 levels were significantly higher than in healthy donors (p<0.001); Supplementary Figure 4. Serum sST2 levels at admission were higher in males (males 59.6[38] ng/mL vs. females 45.4[26]; [p=0.012]; Supplementary Figure 3). There were no differences in sST2 concentrations across age groups at any sampling time (Supplementary Figure 2).

In COVID-19 patients sST2 concentrations peaked at 48-72h control measurement, representing on average a 150% increase over admission (53.1 ng/mL [IQR: 30.9] admission vs 79.3 [IQR: 64.2] control; p <0.001). sST2 concentrations at discharge had significantly declined (44.9 ng/mL [IQR: 39.6]; p<0.001), but were still significantly above healthy donor’s levels; Supplementary Figure 4.

Main characteristics of the cohort stratified according to admission sST2 tertiles are shown in Table 1. Patients in the upper tertile for sST2 at admission (> percentile 75; cut-off=70.4 ng/mL), tended to be male in a higher proportion (p=0.066), and showed no differences for comorbidities. Patients in this P>75 group had worse respiratory function as assessed by estimated PAFI (p<0.001), higher concentrations of aspartate aminotransferase (AST) (p<0.001), alanine aminotransferase (ALT) (p=0.006), creatin-phosphokinase (CK) (p=0.007), lactate dehydrogenase (LDH) (p≤0.001), C-reactive protein (p=0.005), ferritin (p=0.007), D-Dimer (p=0.030), fibrinogen (p=0.011) and interleukine-6 (p=0.008), and higher total lymphocyte count (p=0.021). There were no differences in treatment schedule at admission.

Admission sST2 concentrations showed a significant negative correlation with estimated PAFI (r= − 0.361; p≤ 0.001), and significant positive correlations with LDH (r=0.328; p<0.001), C-reactive protein (r=0.274; p=0.001) and IL-6 serum concentrations (r=0.271; p≤ 0.001).

### Association between sST2 concentrations and outcomes

Fifteen out of 144 (10.4%) patients reached primary endpoint. Fourteen patients required admission to ICU. One patient died of bacteriemia associated to a central venous catheter. Patients with the highest sST2 admission concentrations (> percentile 75), experienced the primary outcome in a higher proportion (P75 = 25.7% vs. P25 = 0%; p= 0.001, Table 2)

**Table 2:**
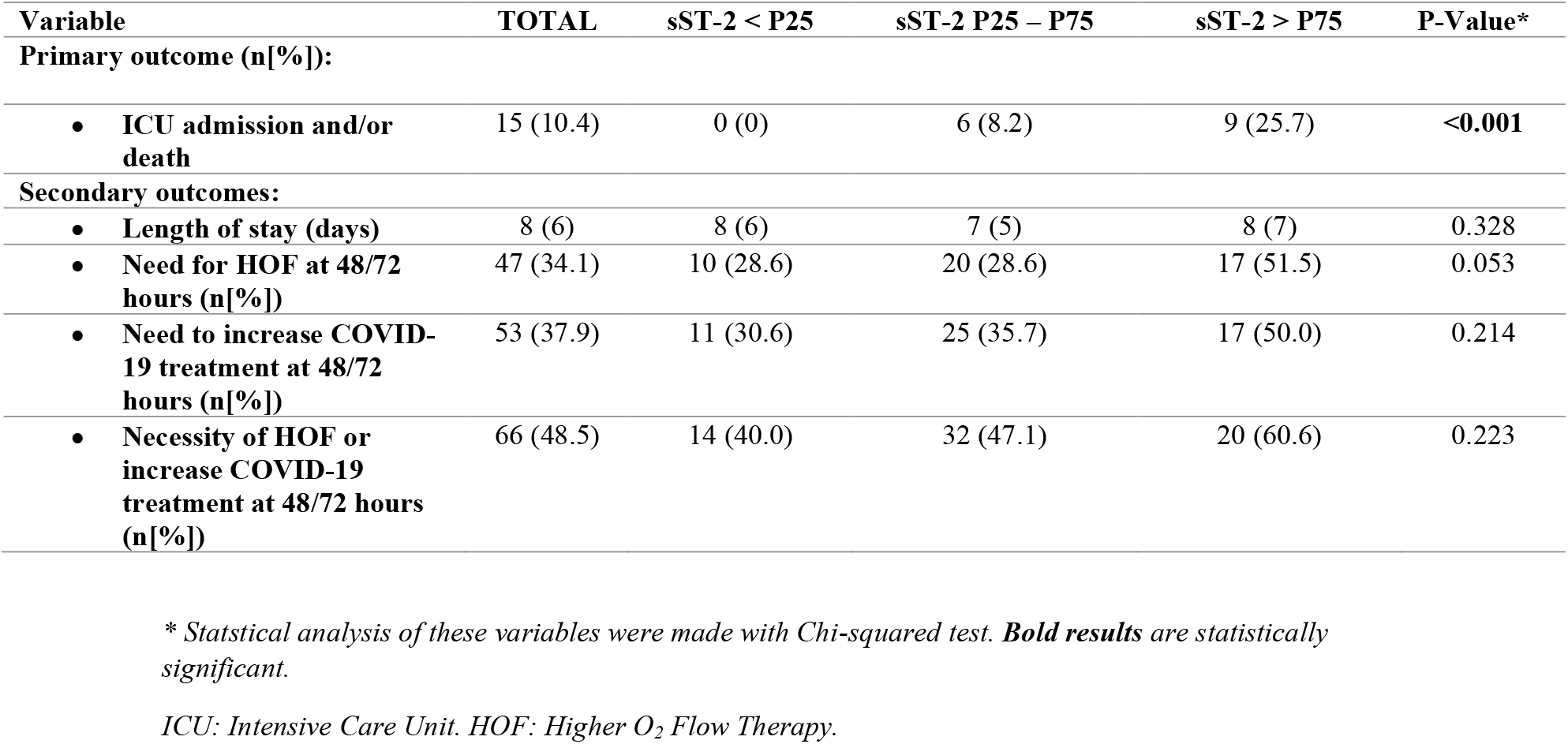
Outcomes by sST-2 concentrations at admission.

Admission sST2 showed the best area under the curve (AUC=0.776; p=0.001) compared with LDH, IL-6, ferritin, CK and C-reactive protein (Figure 1). A cut-off value of 58.9 ng/mL identifies the primary outcome with the best Sensitivity (78.6%) and Specificity (60.8%). Based on this cutoff, Kaplan Meier survival curves for the primary endpoint by sST2 concentrations at admission were generated and showed significantly differences in mortality and ICU admissions (Log-rank test ≤0.001; Figure 2).

**Figure 1:**
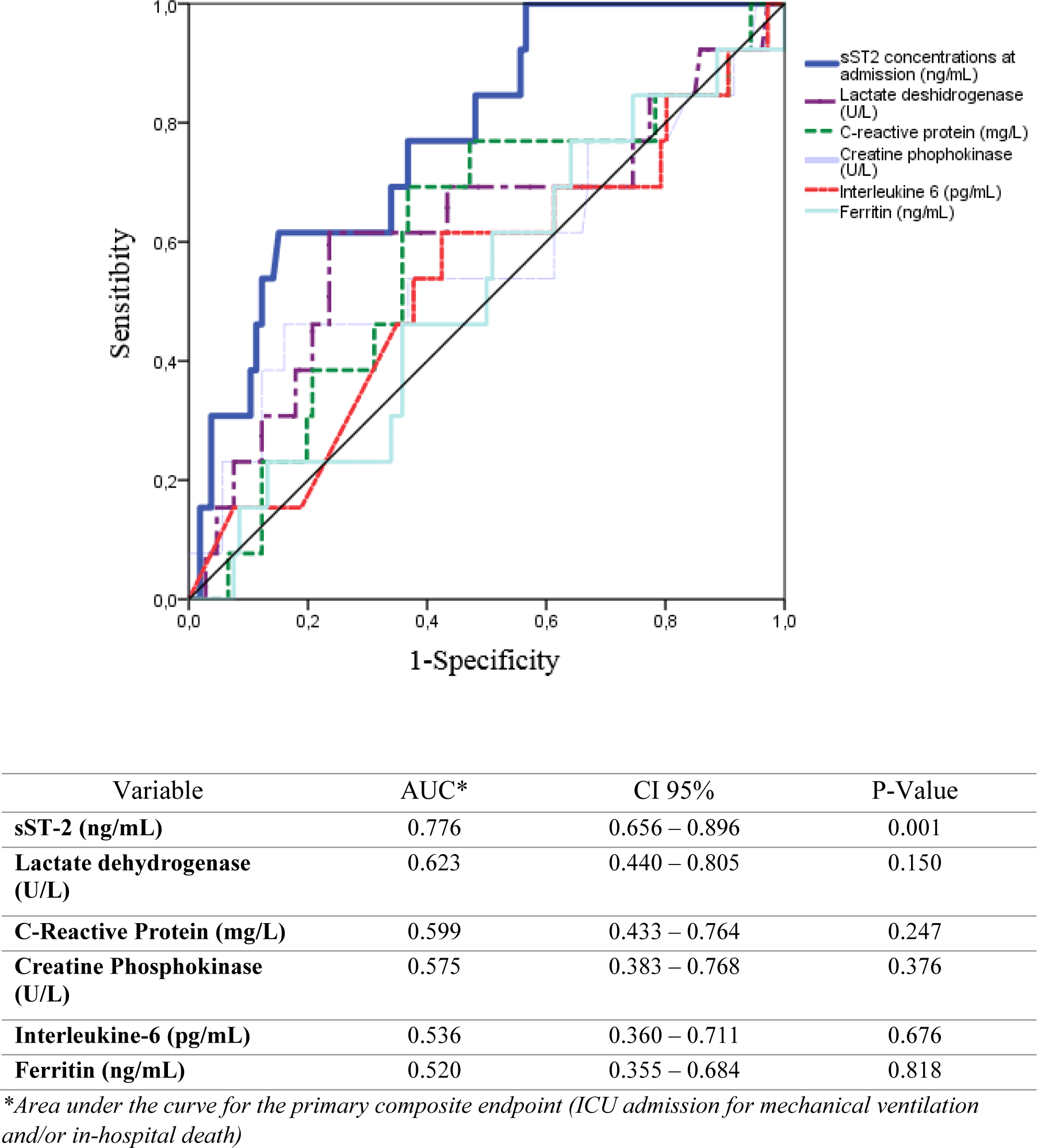
Receiver operating characteristic curves comparing sST2 concentrations at admission with C-reactive protein, lactate dehydrogenase, ferritin and interleukine-6 concentrations at admission.

**Figure 2:**
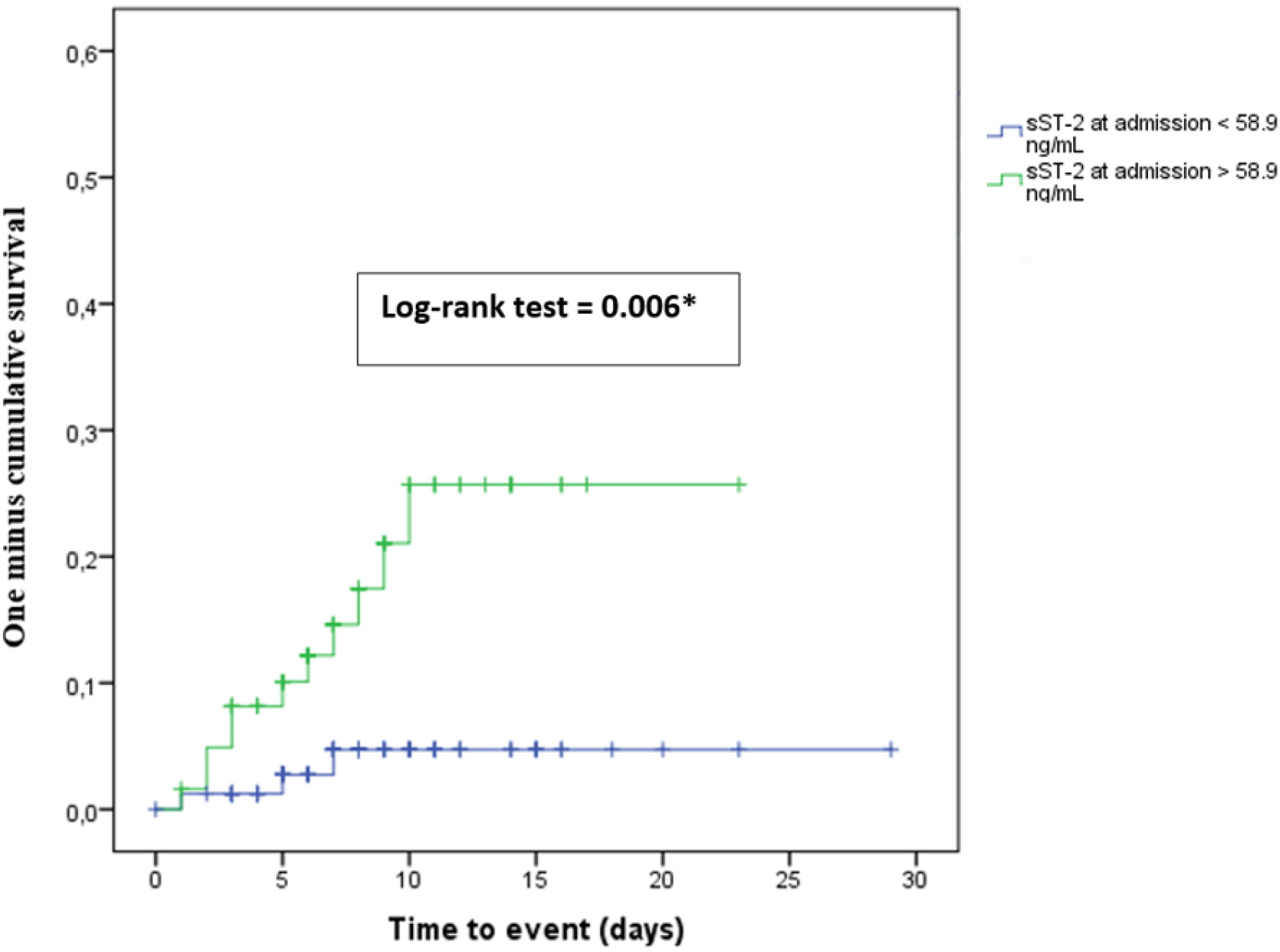
Kaplan Meier curves for the primary outcome (ICU admission for mechanical intubation and/or in-hospital death) by sST-2 concentrations at admission. *Statistical significant

Median length of stay for patients not requiring admission to ICU was 8 days (IQR 6). sST2 values at admission or control time points, or the increase between them, did not correlate with length of hospital stay. However, there was a negative correlation between length of hospital stay and sST2 at discharge (Spearman’s rho = −0.338, p=0.003).

At control time, 47 patients (34.1%) needed HOF, oxygen administration had to be increased in 53 patients (37.9%), and 66 patients (48.5%) required intensification of treatment (Table 2). Proportion of patients suffering these secondary outcomes was higher in the >P75 admission sST2 group for all three events, but only reached significance for HOF (> percentile 75 = 51,5% vs P25 = 28.6%; p=0.053).

Univariable logistic regression analysis identified ePAFI (HR 0.98 [0.97 – 0.99]; p=0.001), CK (HR 2.06 [1.01 – 4.20]; p= 0.047), and admission sST2 > 58.9 ng/mL (HR 6.32 [1.70 – 23.5]; p=0.006), as potential predictors for primary end-point.

In the multivariable logistic regression model, after adjusting for confounders, including age, gender and previous history of diabetes, admission sST2 concentrations (HR 4.53 [1.15 – 17.8]; p=0.031) together with ePAFI (HR 0.04 [0-0.53]; p=0.014) were identified as independent predictors for the primary endpoint (Table 3). AUC of the multivariable model improved significantly when sST2 admission concentrations were included (0.71 [0.567-0.871] vs. 0.81 [0.72 – 0.90]). (Table 3)

**Table 3:** Univariable and multivariable logistic regression model for the primary combined endpoint all-cause mortality and/or ICU admissions.

| Variable | UNIVARIABLE |  | MULTIVARIABLE |  |
| --- | --- | --- | --- | --- |
|  | HR (CI 95%) | P-Value | HR (CI 95%) | P-Value |
| Age | 1.04 (0.99 – 1.09) | 0.073 |  |  |
| Gender-male | 1.38 (0.47 – 4.05) | 0.555 |  |  |
| Diabetes | 2.73 (0.84 – 8.82) | 0.094 |  |  |
| SBP | 1.00 (0.97 – 1.03) | 0.937 |  |  |
| DBP | 0.97 (0.93 – 1.03) | 0.366 |  |  |
| Estimated PAFI* | 0.98 (0.97 – 0.99) | **0.001** | 0.04 (0.00 – 0.53) | **0.014** |
| Urea* | 1.45 (0.55 – 3.83) | 0.455 |  |  |
| Aspartate transaminase * | 1.31 (0.51 – 3.41) | 0.576 |  |  |
| Alanine transaminase* | 0.94 (0.41 – 2.15) | 0.941 |  |  |
| Creatin phosphokinase* | 2.06 (1.01 – 4.20) | **0.047** |  |  |
| Lactate deshydrogenase* | 5.13 (0.93 – 28.2) | 0.060 |  |  |
| C-Reactive Protein* | 1.33 (0.73 – 2.44) | 0.357 |  |  |
| Ferritin* | 1.00 (0.56 – 1.80) | 0.999 |  |  |
| Total lymphocytes* | 1.13 (0.54 – 2.36) | 0.747 |  |  |
| D-Dimer* | 1.23 (0.68 – 2.22) | 0.501 |  |  |
| Fibrinogen* | 0.57 (0.06 – 5.51) | 0.629 |  |  |
| Interleukine-6* | 1.28 (0.63 – 2.58) | 0.491 |  |  |
| sST2 (cut-off > 58.9 ng/mL †) | 6.32 (1.70 – 23.5) | **0.006** | 4.53 (1.15 – 17.8) | **0.031** |

CPK: Creatin Phosphokinase; DBP: Diastolic blood pressure; SBP: Systolic blood pressure.
List of candidate variables included in multivariable logistic regression model (backward selection procedure): Age, Gender, Previous history of diabetes, Estimated ePAFI, creatin phosphokinase, Lactate deshydrogenase. AUC of the model without sST2: 0.71 [0.567-0.871], AUC of the model: 0.81 (CI95% [0.72 – 0.90]; $p < 0.001$ ).
† Point of highest sensibility and specificity (Sensitivity = 78.6%; Specificity = 60.8%; AUC = 0.776; $p = 0.001$ ).
\* Variables have been transformed using logarithmic polynomials

## Discussion

Here we present our results demonstrating that early changes in blood sST2 have an excellent prognostic value in COVID-19, surpassing more traditional markers having been used during the pandemic. An added value to our findings is the fact they have been produced from a cohort of patients managed under usual clinical practice conditions.

Emergence of COVID-19 has posed an unprecedent challenge for clinicians around the World. COVID-19 exhibits a wide clinical spectrum ranging from asymptomatic or mild respiratory tract symptoms, to the development of an ARDS and death. Respiratory failure is usually the hallmark of bad prognosis and mortality^1,2^. The reasons why patients, presenting with apparently similar clinical features, evolve to a severe disease or keep mildly affected remain to be fully elucidated. But worse, development of severe lung affectation has proven difficult to predict. Moreover, COVID-19 has put significant strain on healthcare systems, and tools assisting in evidence-guided decision making to allocate limited resources are in urgent need.

In an attempt to anticipate clinical course, a number of diagnostic tests and biomarkers have been evaluated and are currently the standard approach. Among the biomarkers, there are classic indicators of cell damage (LDH, troponin) and biomolecules suspected to be involved in the immune and inflammatory response firstly identified in disease pathogenesis (IL-6, ferritin or lymphocytes count) ^1,3,15^. None of them are specific, which complicates risk stratification in a disease following worse course in patients with chronic comorbidities characterized by low-degree inflammation.

We found that admission sST2 values correlated well with biochemical markers and indexes being used to evaluate clinical course. Multivariate regression models and survival analysis demonstrated that admission sST2 is an independent predictor of worse prognosis in patients admitted for COVID-19. In ROC analysis sensitivity and specificity of sST2 to predict worse outcomes was significantly higher than that of the aforementioned biomarkers. sST2 also correlated to prescription of HOF, transfer to ICU, length of hospital stay and death. Altogether, our findings support the clinical usefulness of sST2 in current clinical setting.

A powerful approach to identify better prognostic biomarkers is to perform unbiased searches for biomolecules exhibiting altered expression in COVID-19. Applying this approach, *Huang* examined the plasma protein profile of COVID-19 patients and the overlaps with common comorbidities (reported in preprint format). Clinical evolution was estimated from longitudinal analysis of WHO 6-point ordinal scale. Baseline sST2 levels were associated with baseline disease severity and with worse outcomes (death or requiring ventilation in 28 days follow-up). sST2 values peaked at day 3^16^. Because the IL-33/ST2 axis is involved in immune response to viral infections, *Zeng et al*. measured IL-33 and sST2 in 80 COVID-19 patients (reported in preprint format). In line with our results, sST2 levels were significantly elevated in COVID-19 and correlated with other markers of inflammation and disease clinical severity. However this study did not address hard clinical endpoints like ICU admission or death^17^. Interestingly, this study did not find significant differences in serum IL-33 between healthy controls and COVID-19 patients. However, a significant association for IL-33 levels at admission with adverse outcome (ICU admission, requiring ventilation or death) has been reported, with a very good predictive value (AUROC 0.83) in patients <70 years^18^. Another recent reprint looked for cytokine signature and prognosis association in 175 COVID-19 patients. sST2 was found, together with other 7 markers, to be independently associated with mortality both at baseline and longitudinally^19^. Thus, although scarce and mostly in preprint format, available evidence supports our finding that sST-2 is elevated in COVID-19 and that such increase bears prognostic significance.

A unique aspect of our study is the demonstration that early changes predict outcome better than peak values, stressing their clinical usefulness in prognosis. Moreover, sST2 showed a decline in patients at discharge, displaying a fully dynamic behavior that is not present with other biomarkers. Interestingly, discharge sST2 correlated negatively with hospital stay length. This is suggestive that decline in sST2 is delayed with regard to clinical improvement. Other reports have shown persistently elevated sST2 levels in severe COVID-19 cases as long as 30 days after clinical onset^17,19^. It remains to be elucidated whether sST2 will recover completely to normal values and whether persistent sST2 levels would associate to clinical sequalae in COVID-19 patients.

The course of temporal changes in circulating sST2 might also be informative about disease pathophysiology.

Mounting evidence suggests IL-33/ST2 axis might play a leading role in COVID-19 pathogenesis. Based on data from studies of local lung inflammation, cytokine/immune cell profiling and systemic responses, *Zizzo et al*. hypothesized IL-33 has a pivotal role in immune and inflammatory response to SARS-CoV2 infection. The release of IL-33 by pneumocytes and endothelium, in response to cell damage caused by viral infection, triggers a cascade of events in fibroblasts, alveolar membrane, coagulation and immune system^20^.

IL-33 effects would be auto-amplified by IL-33 induction of IL-33 release and ST2 expression and sST2 release, as observed recently for asthma^21^. IL-33 is a well-known factor in asthma, one of the several lung diseases where IL-33 plays a relevant role^21^. More importantly, IL-33 has also been shown to participate in lung epithelium response to viral infection^22^ and to decrease antiviral innate immunity in this organ^23^. Due to technical difficulties in measuring IL-33 levels, circulating sST2 has been often employed as a surrogate marker of IL-33/ST2 axis status. Several studies have shown serum sST2 concentrations are elevated, and correlate with prognosis, in patients with pulmonary disorders including asthma, idiopathic pulmonary fibrosis, severe sepsis, and trauma ^13^. Bajwa et al. compared sST2 concentrations in patients with ARDS vs. congestive HF. They found higher sST2 concentrations in ARDS, exhibiting good correlation with mortality and APACHE III scale; to conclude sST2 could be a very good biomarker to distinguish ARDS from cardiogenic edema^14^.

Several organs and tissues other than the lung and immune cells, namely the heart, adipose tissue, endothelium, express locally the IL-33/ST2 axis and may release sST2 ^10,9^.Our finding may also support an important role for IL-33/ST2 axis in COVID-19 systemic manifestations. Kawasaki disease is a severe multisystem vasculitis mainly encountered in children and young people. Most experts consider it to be consequence of an anormal immunological response evoked by one or more infectious agents^24^. *Sato et al*. investigated the usefulness of sST2 as a surrogate inflammatory biomarker in Kawasaki disease^25^. They tested sST2 along with other classic biomarkers and showed that troponin and natriuretic peptide were not useful, while sST2 showed high concentrations in patients with Kawasaki disease which, in addition, correlated with clinical worse course^25,,26^. Of note, a rare, but severe pediatric manifestation in COVID-19 has been described which closely resembles Kawasaki disease^27^.

sST2 is also an established prognosis maker in heart failure (HF). sST2 blood concentrations correlate with mortality, alone or in combination with other classical biomarkers such as natriuretic peptides^28^ Interestingly, the study by *Pascual-Figal et al*. provided strong evidence that vascular and pulmonary endothelium, as well as pneumocytes are main sources of ST2 in ADHF^10,9^. The potential association of elevated sST2 levels in COVID-19 with myocarditis signs and markers observed in these patients deserves further consideration, as does the possible consequences for the cardiovascular system of a potentially sustained elevation in sST2 in convalescent patients, identified in this and other studies.

Our study has some limitations. It has been conducted in a single center and on a small sample size, this latter driven by the urgency to advance our knowledge on the disease. Nonetheless, clinical characteristics in our cohort were concordant with published data from other studies around the world and in our country^1,2^. Due to resource limitations imposed by the demanding sanitary situation, access to imaging and functional tests was difficult to standardize for even a representative fraction of study patients and thus their clinical value could not be compared directly to that of sST2. Clinical data for the study was collected under conditions of real practice.

Obviously, this may imply unforeseen bias have been introduced, but multivariable analysis diminished this risk. And in the other hand, in a such heterogenous syndrome and with pandemic assistance, this setting could be more a strength than a limitation.

Identification of sST2 as an early predictor of worst prognosis in COVID-19 patients has important clinical implications. sST-2 is a readily available biomarker, which can be consistently determined in blood samples. Accumulating evidence supports sST2 levels elevate early in SARS-CoV2 infection and that such changes represent an independent risk factor for worse outcome, outpowering other biomarkers that so far have been guiding clinical and therapeutical handling. Another consistent report is the persistence of elevated sST2 after apparent clinical resolution. Given the well-established associations of sST2 levels with other lung and cardiovascular diseases, it is mandatory to investigate whether such sST2 levels reflect some inflammatory memory with long-term consequences. Our findings also lend support to the hypothesis of an IL-33/ST2 axis led pathophysiology in COVID-19. Pharmacological modulators for this axis are available and currently being trialed for other pathologies, that could be evaluated for treating or preventing severe cases of COVID-19.

Among patients suffering from COVID-19 infection, severely enough to be admitted, measurement of sST2 within 24h of admission, may be a useful biomarker for an early identification of those at higher risk of severe complications or death and for implementing more aggressive therapies since the initial stages of the disease.

## Data Availability

All data referred to in the manuscript are guarding by the authors in an anonimyzed database according to National Law of protection o personal data.

## Acknowledgments

To all the staff, physicians, nurses and technicians, of the Internal Medicine and Infectious Diseases Department. To patients who agreed to participate in the study. To all of the patients who suffered and died from the pandemic and health workers who looked after them. And finally, to our families who support and help us in these very difficult moments.

## Conflict of interest

Authors do not declare conflict of interests

## Funding

The study was funded through a COVID-19 2020 crowdfunding campaign launched by the Aragon Health Research Institute (https://www.iisaragon.es/utilidad-de-la-ecografia-clinica-y-el-uso-de-biomarcadores-sericos-en-la-estratificacion-del-riesgo-de-pacientes-con-infeccion-por-sars-cov2-covid-19/).

## Notes

### Competing Interest Statement

The authors have declared no competing interest.

### Author Declarations

The study complied with the fundamental guidelines of the Helsinki declaration guidelines and was evaluated and approved by Aragon's Committee on Research Ethics (CEICA, Ref. PI20/248, May 13Th, 2020).

