## SUPPLEMENTARY MATERIAL for "Early measurement of blood sST2 is a good predictor of death and poor outcomes in patients admitted for COVID-19 infection"

**Supplementary material: ST2 circulating measurements details**

Serum sST2 concentrations were determined in 150 COVID-19 patients admitted at hospital. Blood samples were withdrawn at admission, control and discharge. Blood was collected into clotting gel test tubes, centrifuged and serum was aliquoted and stored (Aragón's Health System Biobank) at -80ºC until analysis. All biological samples were collected under informed consent. Eventually, 144 admission and control, and 80 discharge samples were processed and analyzed. Serum aliquots were virus-inactivated by treatment with 1% Triton-X100. On the day of the analysis, serum was thawed and diluted 1:50 in Reagent Diluent buffer (1% Bovine Serum Albumin in Phosphate Buffered Saline). Serum concentrations of soluble ST2 were determined by sandwich enzyme-linked immuno-sorbent assay (ELISA), and following instructions provided by kit manufacturer (DY523B, R&D Systems Europe Ltd). Briefly, serum samples and standards were incubated with capture antibody adsorbed to 96 well plates, followed by incubation with detection antibody. Analyte-antibody complexes were detected using a Horseradish Peroxidase-based colorimetric assay and OD450 signal was recorded using a microplate reader. sST2 serum concentrations were calculated from standard curve after blank subtraction. A set of sera from 60 healthy donors obtained through Aragón's Health System Biobank (BSSA) was also analyzed in this manner. These sera had been originally collected from two independent sources, and were selected to match patient cohort age and gender distribution. Random samples from COVID patients and healthy donors were re-run in independent assays to test and correct for inter-assay variability.

**Supplementary Figure 1:** Inclusion Flow chart

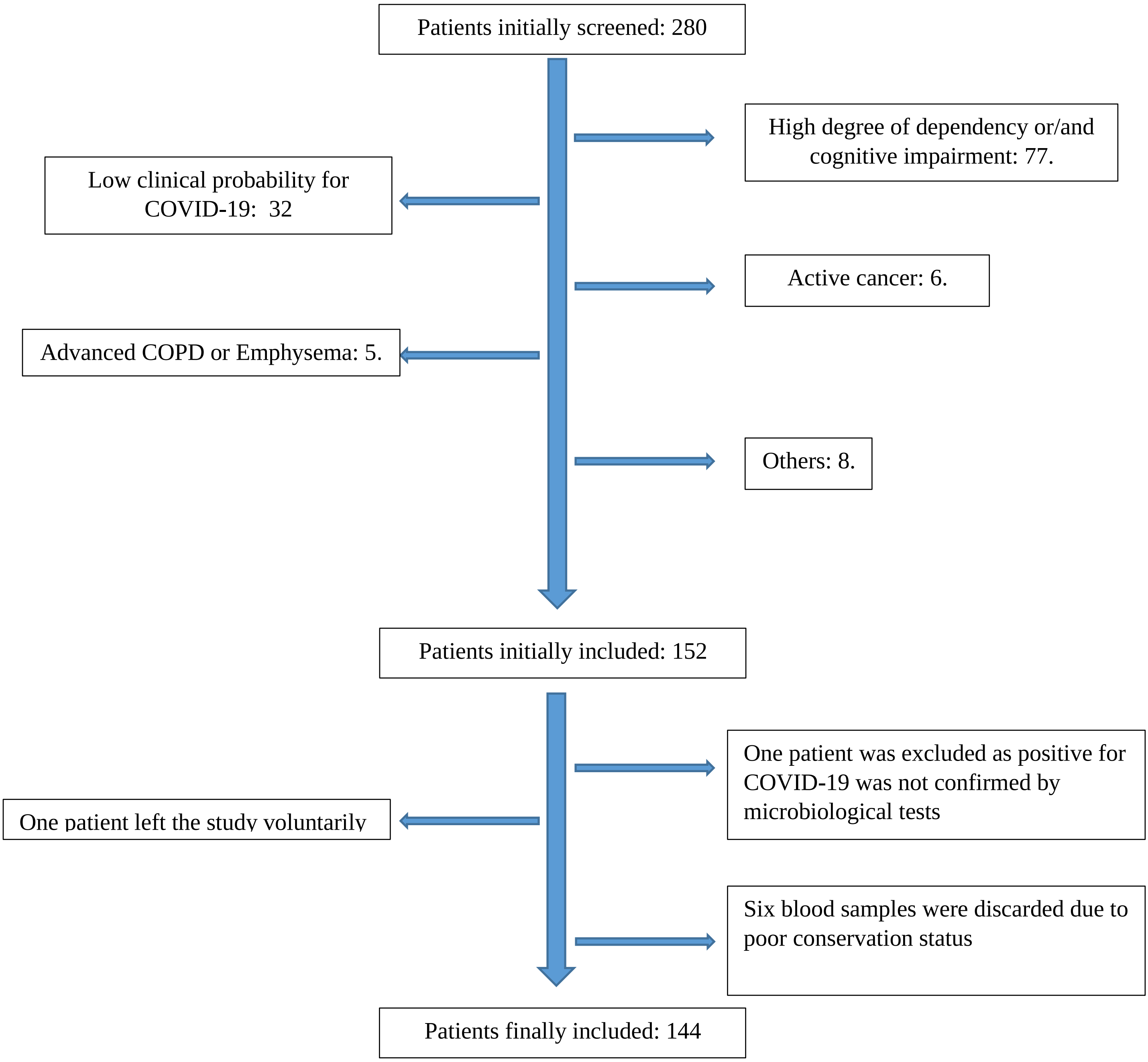

**Supplementary Figure 2:** sST2 concentrations across age groups

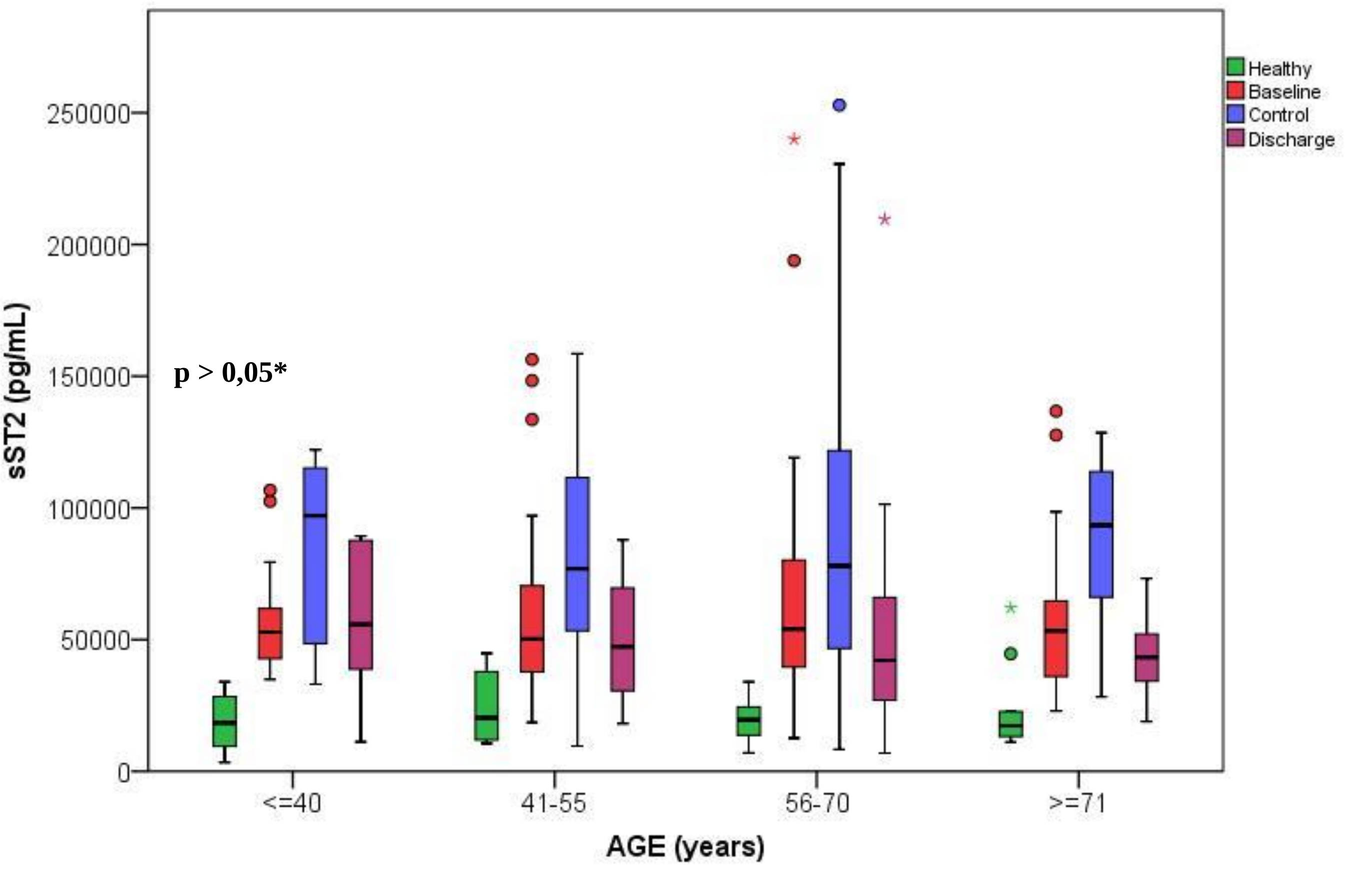


*There were no differences between age groups in healthy donor, neither COVID-19 patients (p>0,05)

**Supplementary Figure 3:** sST2 concentrations in male and female

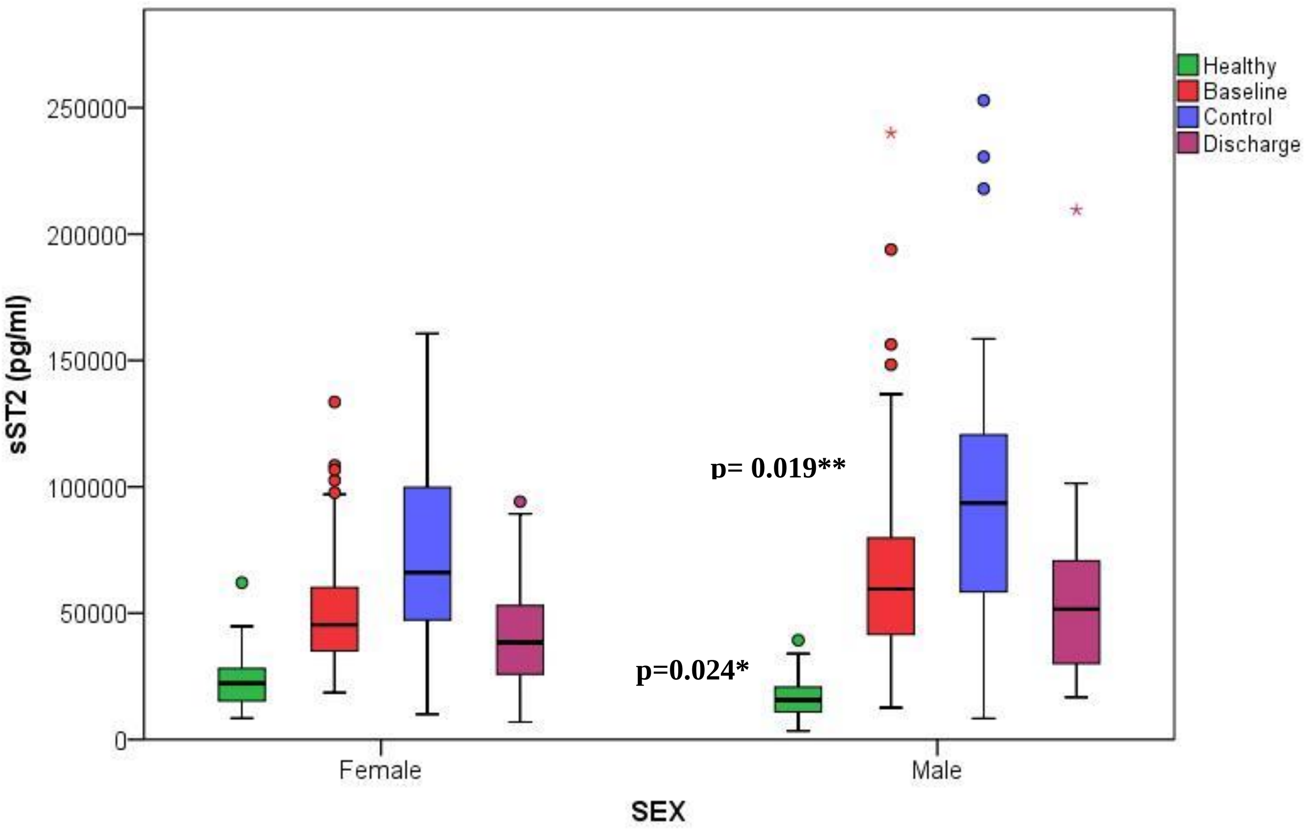


*sST2 concentration was higher in female healthy donors than male (p=0,024)

**sST2 concentration was higher in males vs females (p= 0.012)

**Supplementary Figure 4:** sST2 concentrations in healthy donors and COVID-19 patients

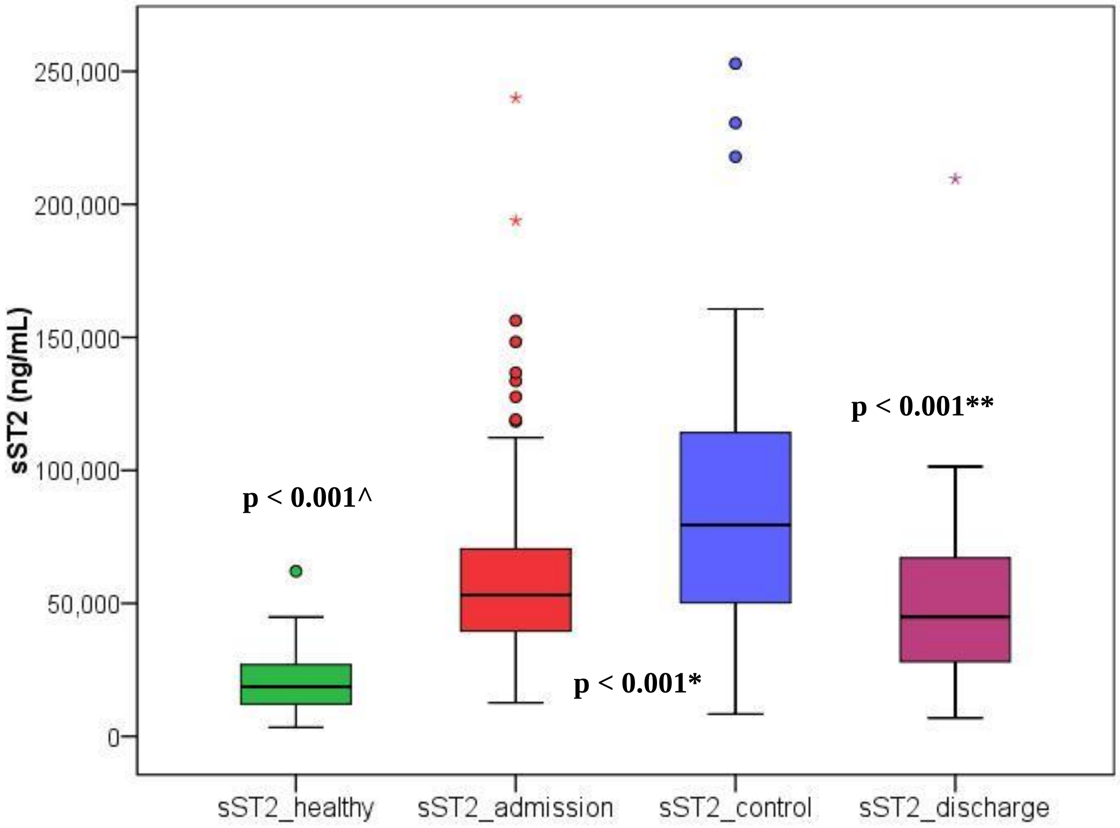


^Wilcoxon test between sST-2 concentrations in healthy donors and COVID-19 patients.
*Wilcoxon test between sST-2 concentrations at admission and sST-2 concentrations at control.
** Wilcoxon test between sST-2 concentrations at control and sST-2 concentrations at discharge (Median time of discharge blood analysis (7 days [5]).
